# Perceptions of Artificial Intelligence in the Editorial and Peer Review Process: A Cross-Sectional Survey of Traditional, Complementary, and Integrative Medicine Journal Editors

**DOI:** 10.64898/2026.03.04.26347571

**Authors:** Jeremy Y. Ng, Daivat Bhavsar, Malvika Krishnamurthy, Neha Dhanvanthry, Daniel Fry, Ji Woo Kim, Amelia King, Jaimie Lai, Anthony Makwanda, Priscilla Olugbemiro, Jeel Patel, Insha Virani, Ella Ying, Kingsley Yong, Abdullah Zaidi, Jasmine Zouhair, Myeong Soo Lee, Ye-Seul Lee, Tanuja M. Nesari, Thomas Ostermann, Claudia M. Witt, Linda Zhong, Holger Cramer

## Abstract

**Background:** Artificial intelligence chatbots (AICs) are increasingly being integrated into scholarly publishing, with the potential to automate routine editorial tasks and streamline workflows. In traditional, complementary, and integrative medicine (TCIM) publishing, editorial and peer review processes can be particularly complex due to diverse methodologies and culturally embedded knowledge systems, presenting unique opportunities and challenges for AIC adoption.

**Methods:** An anonymous, online cross-sectional survey was distributed to the editorial board members of 115 TCIM journals. The survey assessed familiarity and current use of AICs, perceived benefits and challenges, ethical concerns, and anticipated future roles in editorial workflows.

**Results:** Of 5119 invitations, 217 eligible participants completed the survey. While approximately 70% of respondents reported familiarity with AI tools, over 60% had never used AICs for editorial tasks. Editors expressed strongest support for text-focused applications, such as grammar and language checks (81.0%) and plagiarism/ethical screening (67.4%). Most respondents (82.8%) believed that AICs would be important or very important to the future of scholarly publishing; however, the majority (65.3%) reported that their journals lacked AI-specific policies and training programs to guide editors and peer reviewers.

**Conclusions:** Most TCIM editors believe that AICs have potential to support routine editorial functions but also have limited adoption into editorial and peer review processes due to practical, ethical, and institutional barriers. Additional training and guidance are warranted by journals to direct responsible and ethical use if AICs are to be adopted in TCIM academic publishing.

## Background

Artificial intelligence (AI) has been increasingly integrated into various stages of the publishing process, offering promising tools to enhance the efficiency, accuracy, and objectivity of the research process [1]. In the editorial and peer review process, AI applications have the potential to not only support automated manuscript screening, plagiarism detection, recommending reviewers, and providing initial reviews, but also whole processes [2,3]. These technologies aim to streamline workflows, reduce the time to publication, and ensure the integrity of scientific literature.

Traditional, complementary, and integrative medicine (TCIM) is a diverse field encompassing various practices and treatments, differing by cultural contexts in different countries, that are not traditionally part of conventional medical care, such as acupuncture, herbal remedies, and meditative techniques [4]. The editorial and peer review process in TCIM journals can be particularly challenging due to the interdisciplinary nature of the research, the diversity of methodologies, and the differing viewpoints of TCIM researchers, among other stakeholders [5,6]. AI offers unique opportunities to address these challenges by providing tools that can assist in evaluating complex methodologies, ensuring the reproducibility of studies, and maintaining high standards of evidence [2,7,8].

Despite the potential benefits, the adoption of AI in the editorial and peer review process within TCIM journals is not well-documented. There is a paucity of empirical data on how editors perceive the role of AI, its benefits, challenges, and ethical implications in this specific context [9,10]. Understanding these perceptions is crucial for informing the development and implementation of AI tools that are tailored to meet the needs of TCIM journals and for addressing any concerns that may hinder their adoption [10]. This knowledge gap underscores the need for comprehensive research to evaluate the attitudes and experiences of TCIM journal editors regarding the role of AI in the peer review and editorial processes.

Exploring the perceptions of TCIM journal editors regarding AI is timely and significant. AI can potentially enhance efficiency by reducing the workload of editors and reviewers through the automation of routine tasks, allowing them to focus on more critical aspects of the review process [2,7]. Moreover, AI tools can help improve quality and consistency in manuscript evaluations, reducing human biases and enhancing the overall quality of published research [11,12]. Addressing, such as transparency, accountability, and potential biases associated with AI, is essential for gaining the trust of editors and the broader scientific community [1,12,13]. Insights from this study may guide the development of AI tools that are more effectively aligned with the specific needs and values of the TCIM research community.

This study aims to fill the existing knowledge gap by providing a comprehensive assessment of TCIM journal editors’ perceptions of AI in the editorial and peer review process through an anonymous online survey. The survey findings have important implications for the future integration of AI technologies in academic publishing, particularly within the field of TCIM.

## Methods

### Open Science Statement

This study was registered on the Open Science Framework (OSF) and is available here: https://doi.org/10.17605/OSF.IO/UBSJ4. Relevant study materials and data can be found here: https://doi.org/10.17605/OSF.IO/PTFXS. In addition, the full peer-reviewed study protocol has been published [14].

### Study Design

An anonymous, online cross-sectional survey, targeting the editorial board members of 115 journals focusing on TCIM, was adapted from a previous study conducted by Ng et al. [15]. The survey questions were reviewed by subject area content experts (JYN, MSL, YSL, TMN, TO, CMW, LZ, HC).

### Sample and Sampling Method

The inclusion criteria for this study were editors-in-chief (EiCs), associate editors, and other editorial board members of TCIM journals that are directly involved in the peer review and/or editing processes of submitted manuscripts. We used a list of unique TCIM journals (see **Appendix 1**, on OSF at https://osf.io/jz3bm) that was created based on the following search criteria detailed in **Table 1**. For each journal, the names of all editors and editorial board members were collected from the journal webpage. The email addresses of these editors were manually searched and extracted. Editors strictly involved with the manuscript formatting aspects of the editorial process (e.g., copyeditors, technical editors, etc.), statistical editors, and other non-editorial staff that are not responsible for managing the TCIM content contained within manuscript submissions were excluded. Duplicate email addresses were also removed. A total of 14 authors contributed to this task (DB, ND, DF, JWK, AK, JL, AM, PO, JP, IV, EY, KY, AZ, JZ).

**Table 1:**
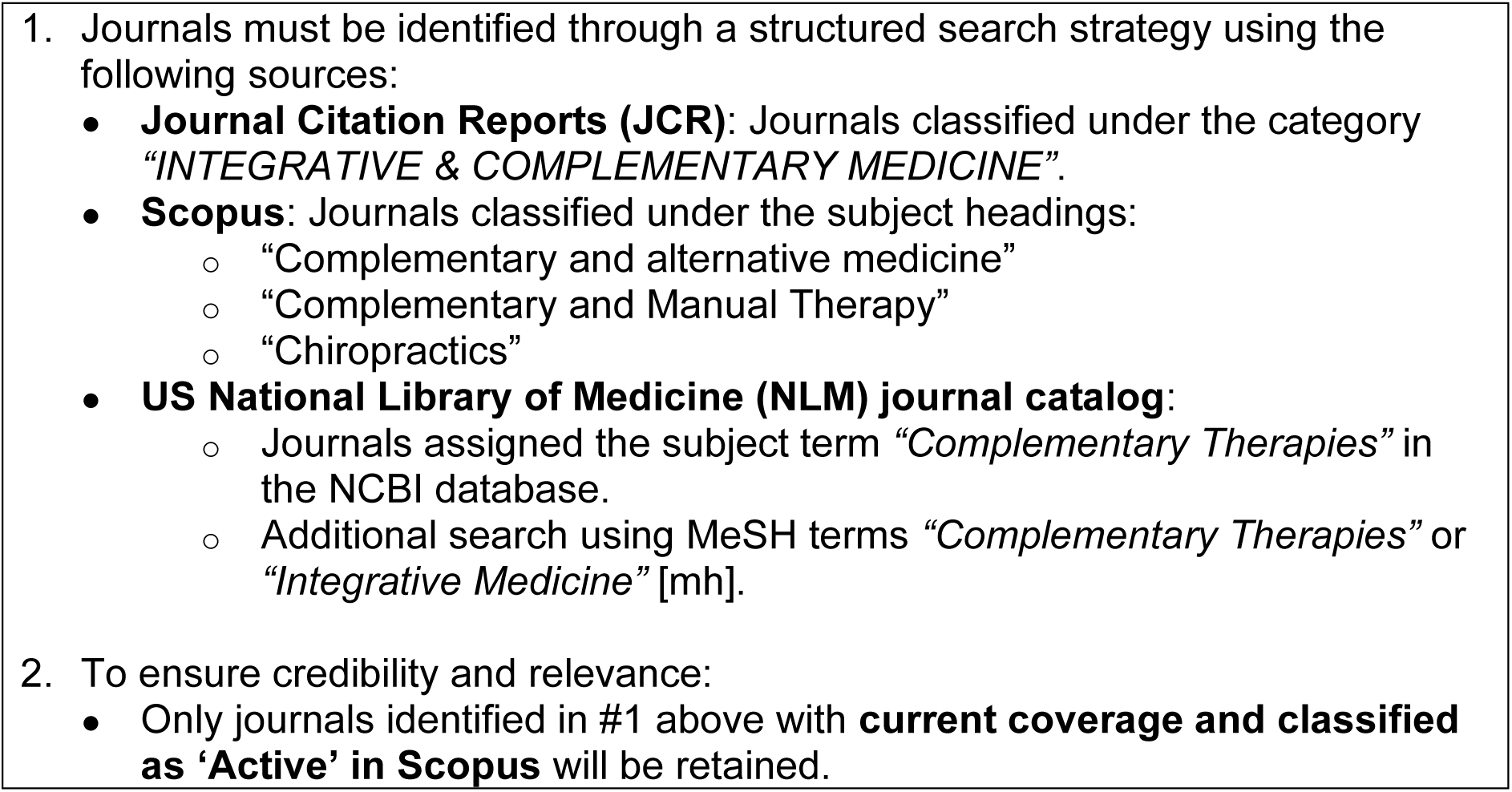
Traditional, Complementary, and Integrative Medicine Journal List Search Criteria.

The survey was closed; only email invitees had access to this survey, and they were instructed not to share the survey link with others. Emails were sent out to the prospective participants via SurveyMonkey [16]. The initial email included a description of the study, the objectives, and a link to access the survey. Clicking on the survey link led the participants to an informed consent form. After the participants provided consent, participants were taken to the survey questions. Numbers of invalid and non-functional emails were recorded by SurveyMonkey. Financial compensation was not provided for participation. Moreover, there was no requirement for those who were emailed the survey link to partake in this study. Withdrawal of responses after survey submission was not possible as survey responses were collected anonymously.

### Survey Development

The survey instrument was published with the study protocol [14]. The survey was open for a total of 6 weeks from May 26, 2025 to July 07, 2025; invitees who had not responded to the original invitation received follow up emails on June 02, 2025, June 09, 2025, and June 16, 2025.

The survey contained 38 questions, spanning 9 pages on Microsoft Word, and took approximately 15 minutes for participants to complete. The survey was developed as a structured questionnaire which began with a screening question for eligibility, and featured sections on demographics and background information, current use and familiarity with AI in editorial tasks, perceived benefits and challenges of AI, ethical concerns and potential biases of AI, and perceptions of the future outlook on AI in publishing. Participants had the option to skip any questions (apart from the screening question) they did not wish to answer, and no personal identifying information was collected. The questionnaire underwent pilot testing with a small group of participants with editorial expertise in the field of TCIM, and revisions were incorporated as appropriate to improve the face validity of this survey.

### Data Analysis

Descriptive statistics (e.g., ratios, percentages) for demographic data was generated from the analysis of the quantitative survey data. Open-ended responses were analysed using inductive coding and thematic analysis [17,18]. Each response was first assigned a code that captures the core meaning by two authors (DB, MK), independently. A shared codebook was developed through consensus between the two authors (DB, MK), and the codes were then grouped into broader themes based on emerging patterns and commonalities, then reviewed by JYN. An inductive approach, without reliance on pre-existing theories, guided the analysis to remain grounded in the data itself [17, 18]. The Checklist for Reporting Results of Internet E-Surveys (CHERRIES) and STrengthening the Reporting of OBservational studies in Epidemiology (STROBE) were used to inform the reporting of this survey study [19,20].

## Results

### Section 1: Respondent Demographics (Q1-14)

A total of 5119 survey invitations were sent out, and 579 emails bounced (i.e., survey invitation did not reach the participant). Responses were provided by 237 invitees (5.2% response rate), wherein 217 respondents met the eligibility criteria (91.6%). There is a variable number of responses for each question, as some respondents did not complete all survey questions. The raw survey data with all personal identifiers redacted is available in **Appendix 2** on OSF at https://osf.io/7faem.

Demographic data is presented in **Table 2**. The majority (54.9%) of participants identified as male (n=106/193), and 42.5% identified as female (n=82/193), the remaining participants preferred not to say or chose to self-describe (n=5/193, 2.5%). The top 3 age ranges of participants were: 46-55 years (n=51/193, 26.4%), 65+ years (n=50/193, 25.9%), and 56-65 years (n=44/193, 22.8%). The majority of respondents identified as not being part of a visible minority group (n=148/193, 76.7%) and not having a disability (n=182/193, 94.3%). Respondents were asked where they were located based on the World Health Organization World Regions with the top three regions being the Americas (n=78/193, 40.4%), Europe (n=45/193, 23.3%), and South-East Asia (n=28/193, 15.0%). Respondents were largely editorial board members (n=146/193) or associate/executive editors (n=61/193, 31.6%), faculty members at a university/academic institution (n=135/193, 70.0%), and primarily edited clinical research manuscripts (n=95/193, 49.2%).

**Table 2.** TCIM Editor Demographics.

| **Sex** | **n (%)** |
| --- | --- |
| Male | 106 (54.9) |
| Female | 82 (42.5) |
| Prefer not to say | 4 (2.1) |
| Prefer to self-describe | 1 (0.5) |
| **Age** |  |
| 18-24 | 0 (0.0) |
| 25-35 | 9 (4.7) |
| 36-45 | 33 (17.1) |
| 46-55 | 51 (26.4) |
| 56-65 | 44 (22.8) |
| 65+ | 50 (25.9) |
| Prefer not to say | 6 (3.1) |
| **Part of a Visible Minority Group** |  |
| Yes | 25 (12.9) |
| No | 148 (76.7) |
| Not sure | 12 (6.2) |
| Prefer not to say | 8 (4.2) |
| **Self-identify as Having a Disability** |  |
| Yes | 9 (4.7) |
| No | 182 (94.3) |
| Prefer not to say | 2 (1.0) |
| **Location Based on World Health Organization World Region** |  |
| Africa | 14 (7.3) |
| Americas | 78 (40.4) |
| Eastern Mediterranean | 8 (4.2) |
| Europe | 45 (23.3) |
| South-East Asia | 29 (15.0) |
| Western Pacific | 19 (9.8) |
| **Current Editorial Board Role(s) in any Journal** |  |
| Editor-in-Chief (or e.g. equivalent role) | 31 (16.1) |
| Associate/executive editor | 61 (31.6) |
| Managing editor | 12 (6.2) |
| Section editor | 24 (12.4) |
| Editorial board member | 146 (75.7) |
| Other | 18 (9.3) |

| **Current Editorial Board Role(s) in TCIM Journal(s)** |  |
| --- | --- |
| Editor-in-Chief (or e.g. equivalent role) | 25 (12.9) |
| Associate/executive editor | 44 (22.8) |
| Managing editor | 9 (4.7) |
| Section editor | 23 (11.9) |
| Editorial board member | 136 (70.5) |
| Other | 26 (13.5) |
| **Current Position** |  |
| Faculty member at a university/academic institution | 135 (69.9) |
| Academic research staff (e.g., research coordinator) at a university/academic institution) | 42 (21.8) |
| Government researcher | 11 (5.7) |
| Clinician/practitioner – conventional (MD, PharmD, etc.) | 22 (11.4) |
| Clinician/practitioner – TCIM (ND, DC, acupuncture, etc.) | 31 (16.1) |
| Pharmaceutical industry/company employee | 4 (2.1) |
| Scholarly communications employee (e.g., scholarly journals/publishing, medical writing) | 4 (2.1) |
| Consultant (e.g., own or employed by research consulting firm) | 14 (7.3) |
| Third sector (E.g., NGO, non-profit) | 8 (4.2) |
| No other roles outside being an editorial board member of a TCIM journal | 4 (2.1) |
| Other | 19 (9.8) |
| **Primary Research Category of Manuscript Submission to Journal** |  |
| Clinical research | 95 (49.2) |
| Preclinical research – in vivo | 16 (8.3) |
| Preclinical research – in vitro | 22 (11.4) |
| Health systems research | 8 (4.2) |
| Health services research | 11 (5.7) |
| Methods research | 9 (4.7) |
| Epidemiological research | 6 (3.1) |
| Other | 26 (13.5) |

### Section 2: Familiarity and Current Use of Artificial Intelligence (AI) (Q15-23, 28-29)

Respondents indicated that they were familiar with AI tools (n=133/190, 70.0%) and previously used AI chatbots such as ChatGPT (n=159/190, 83.7%). However, most respondents had never used AI chatbots for purposes relating to their role as an editorial board member (n=114/187, 61.0%) (**Figure 1**). Respondents were unlikely or very unlikely (n=78/190, 41.1%) or somewhat likely (n=47/190, 24.7%) to use AI tools within the context of their role as an editorial board member in the future.

**Figure 1.**
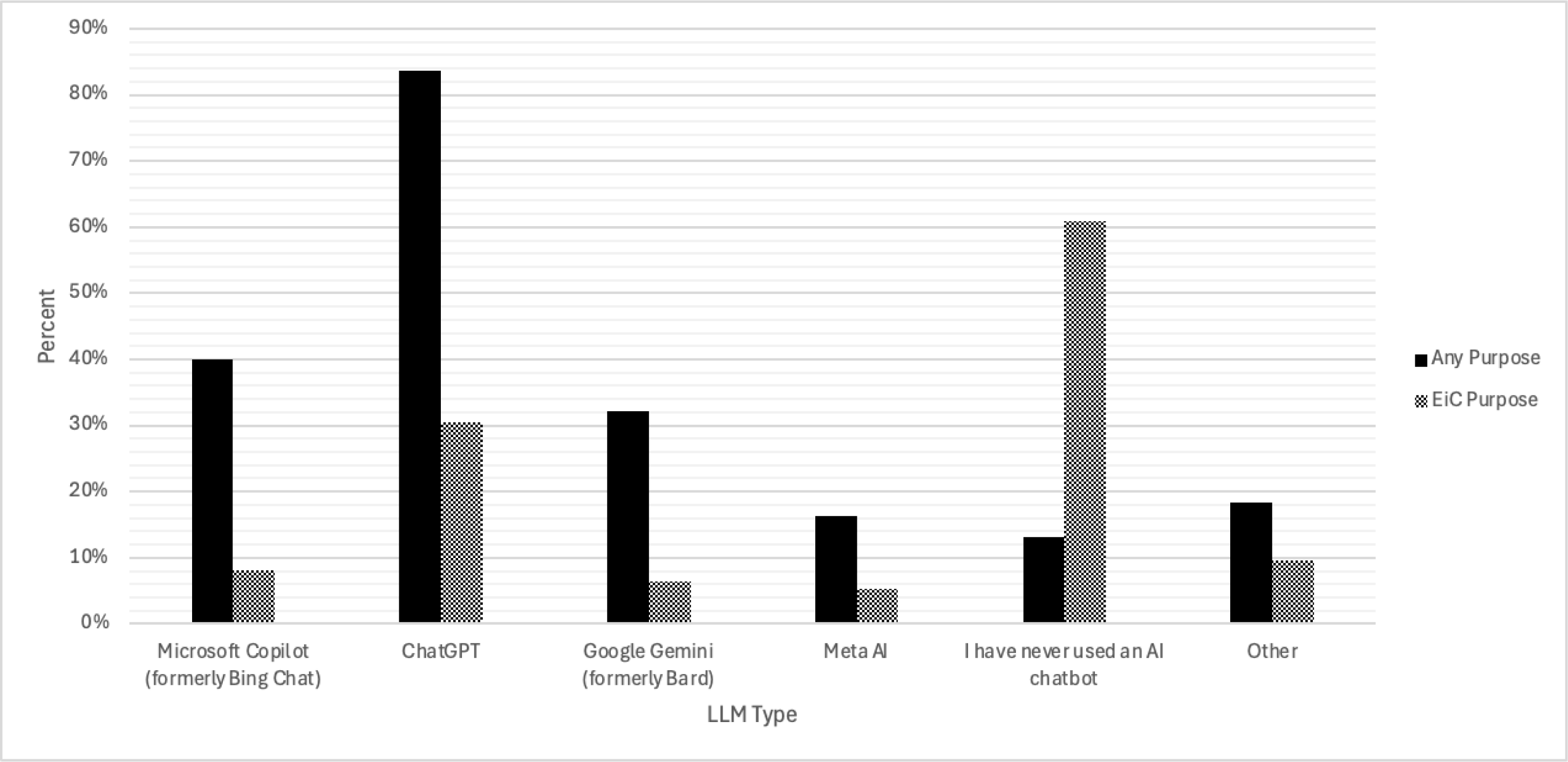
TCIM Editor Use of Various LLMs for Any Use and Editor Specific Use.

Most respondents reported that their affiliated journals or publishers also did not provide any training on AI tools (n=124/190, 65.3%), or that they were not aware if training was provided (n=43/190, 22.6%). Of those who responded “yes” to having journal/publisher affiliated training, most (n=13/24 or 54.1%) responded that they have not taken the provided training. Similarly, when asked if their affiliated journal/publisher has implemented AI tool policies, 30.1% responded “no” (n=57/189) and 46.0% were unsure (n=87/189).

When asked about the amount of training and education that journal editors, including EiCs, would need to effectively use AICs in the editorial and peer review publishing processes, nearly half (n=84/171, 49.1%) indicated “a lot” of training, while 38.6% (n=66/171) believed “some” training would be required. Interest in such training was relatively high, with 63.0% (n=109/173) expressing a willingness to learn more, 23.7% (n=41/173) indicating “maybe,” and only 13.3% (n=23/173) reporting no interest. These results suggest that although most editors perceive a substantial need for training to effectively adopt AICs, many are open to engaging in educational opportunities to build the necessary skills. **Table 3** details the descriptive statistics on the respondents’ familiarity and use of AI, along with available AI training and policies for journal editors.

**Table 3.**
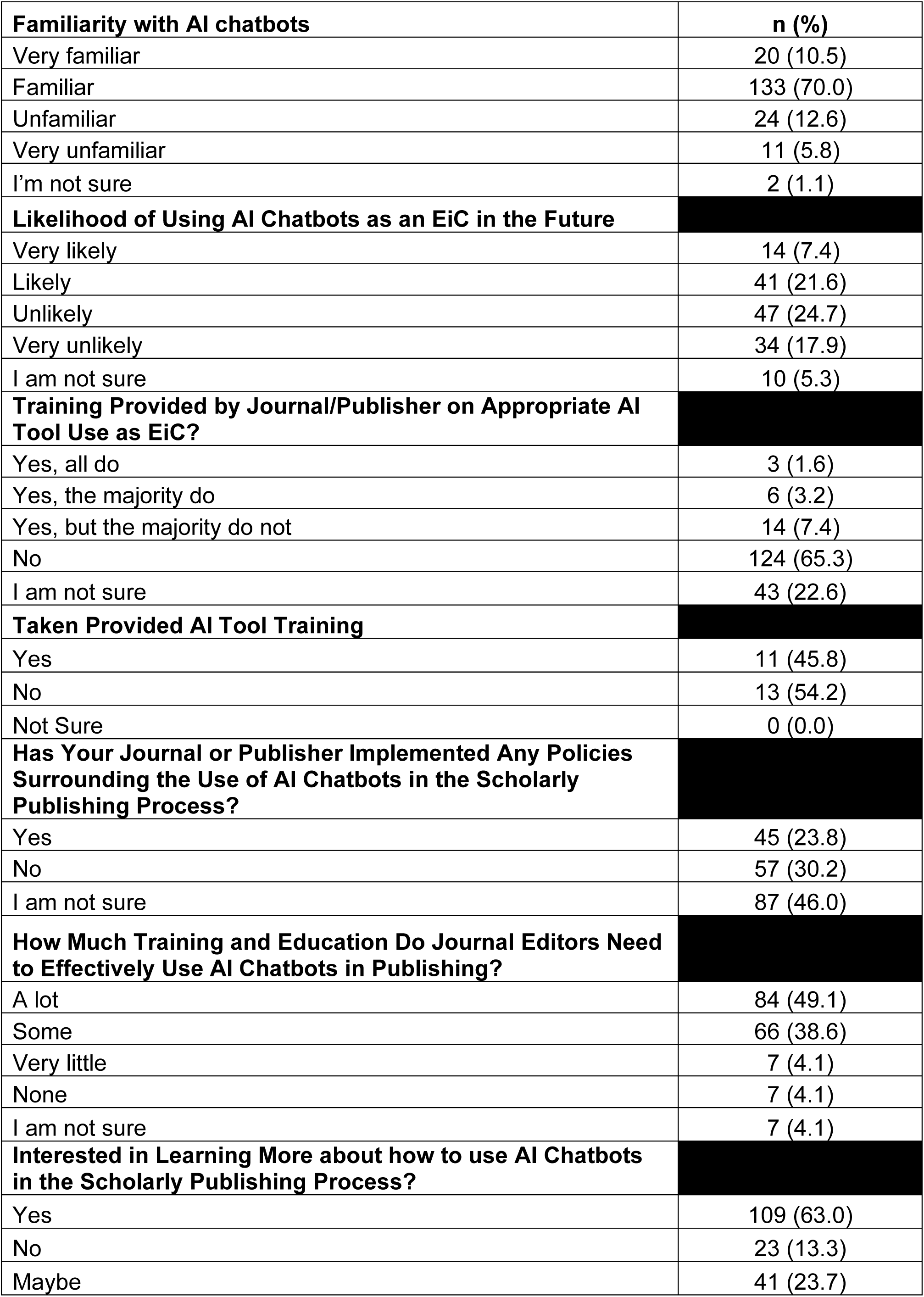
TCIM Editor Familiarity, Use and Training on AICs.

| **Familiarity with AI chatbots** | **n (%)** |
| --- | --- |
| Very familiar | 20 (10.5) |
| Familiar | 133 (70.0) |
| Unfamiliar | 24 (12.6) |
| Very unfamiliar | 11 (5.8) |
| I'm not sure | 2 (1.1) |
| **Likelihood of Using AI Chatbots as an EiC in the Future** |  |
| Very likely | 14 (7.4) |
| Likely | 41 (21.6) |
| Unlikely | 47 (24.7) |
| Very unlikely | 34 (17.9) |
| I am not sure | 10 (5.3) |
| **Training Provided by Journal/Publisher on Appropriate AI Tool Use as EiC?** |  |
| Yes, all do | 3 (1.6) |
| Yes, the majority do | 6 (3.2) |
| Yes, but the majority do not | 14 (7.4) |
| No | 124 (65.3) |
| I am not sure | 43 (22.6) |
| **Taken Provided AI Tool Training** |  |
| Yes | 11 (45.8) |
| No | 13 (54.2) |
| Not Sure | 0 (0.0) |
| **Has Your Journal or Publisher Implemented Any Policies Surrounding the Use of AI Chatbots in the Scholarly Publishing Process?** |  |
| Yes | 45 (23.8) |
| No | 57 (30.2) |
| I am not sure | 87 (46.0) |
| **How Much Training and Education Do Journal Editors Need to Effectively Use AI Chatbots in Publishing?** |  |
| A lot | 84 (49.1) |
| Some | 66 (38.6) |
| Very little | 7 (4.1) |
| None | 7 (4.1) |
| I am not sure | 7 (4.1) |
| **Interested in Learning More about how to use AI Chatbots in the Scholarly Publishing Process?** |  |
| Yes | 109 (63.0) |
| No | 23 (13.3) |
| Maybe | 41 (23.7) |

### Section 3: Specific Roles of AI Chatbots in the Steps of the Scholarly Publishing Process (Q24-33)

Respondents were prompted to rate agreement with statements on a 5-point scale from “very helpful” to “very unhelpful” regarding how helpful they perceive an AIC to be in assisting the different steps of the scholarly publishing process (**Figure 2**). The most favourable perceptions, with respondents indicating either “very helpful” or “helpful,” were observed for language and grammar checks (n=140/172, 81.0%), followed by plagiarism and ethical screening (n=116/172, 67.4%). By contrast, tasks involving interpersonal engagement were met with greater scepticism; handling inquiries from authors, reviewers, and readers was often rated as “unhelpful” or “very unhelpful” (n=67/172, 39.0%), as was direct communication with authors regarding manuscript status and revisions (n=57/172, 33.0%). Process-driven activities attracted comparatively high “not sure” responses; these included monitoring and managing the peer review process (n=61/172, 35.4%), metadata extraction and indexing (n=51/170, 30.0%), and continuous improvement of the publishing workflow (n=61/171, 36.1%).

**Figure 2.**
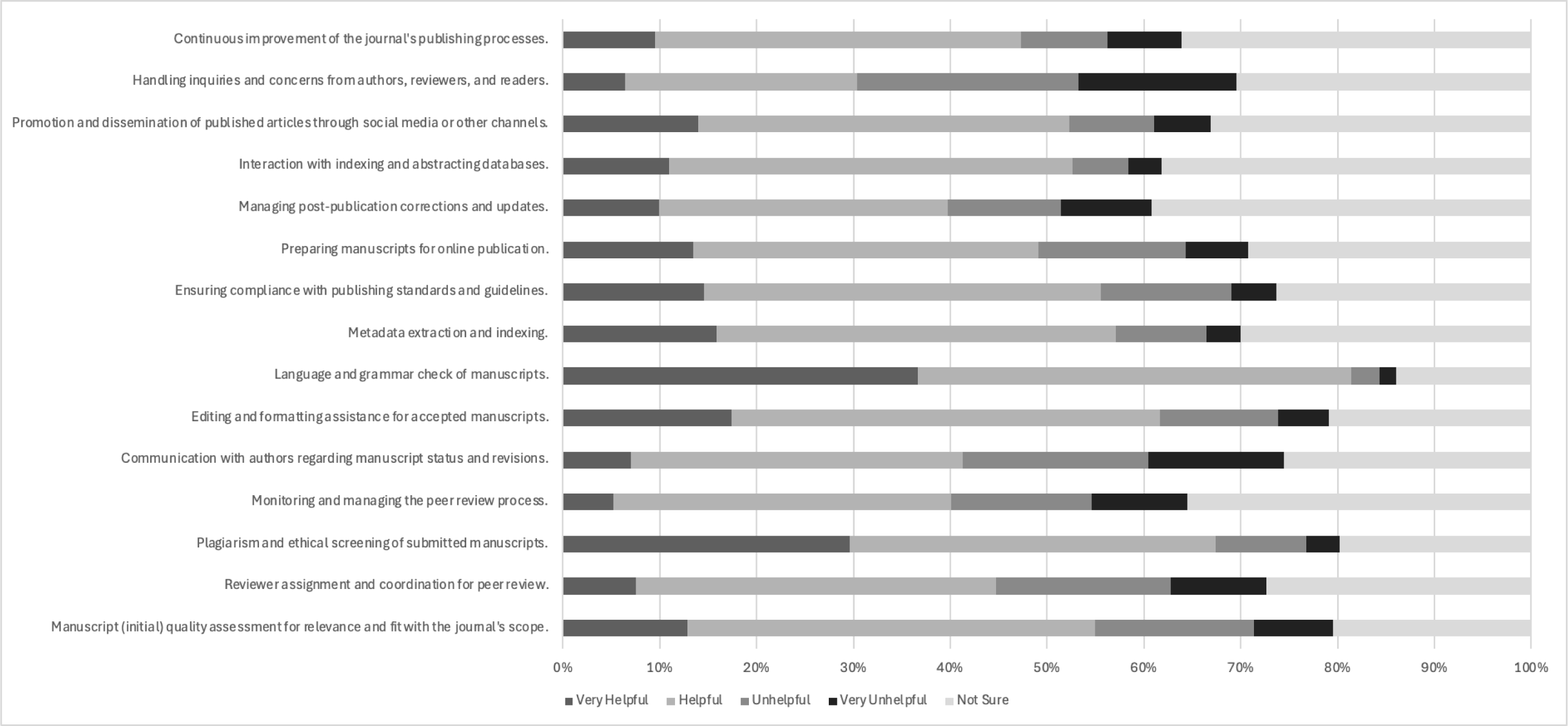
TCIM Editor Perceptions of Steps in Scholarly Publishing Where AICs are Helpful.

Respondents were also asked to indicate how often they had personally used AICs to assist with each step of the scholarly publishing process since the tools became widely available (**Figure 3**). Across all tasks, the most frequent response was “never,” with an average of 71.5% of respondents indicating no AIC use for most publishing activities. The highest rates of non-use were reported for managing post-publication corrections and updates (n=134/171, 78.4%), handling inquiries from authors, reviewers, and readers (n=131/171, 76.6%), and metadata extraction and indexing (n=130/172, 75.6%). More frequent use (“always” or “often”) was reported for language and grammar checks (n=43/172, 25.0%), and plagiarism and ethical screening of submitted manuscripts (n=38/172, 22.1%).

**Figure 3.**
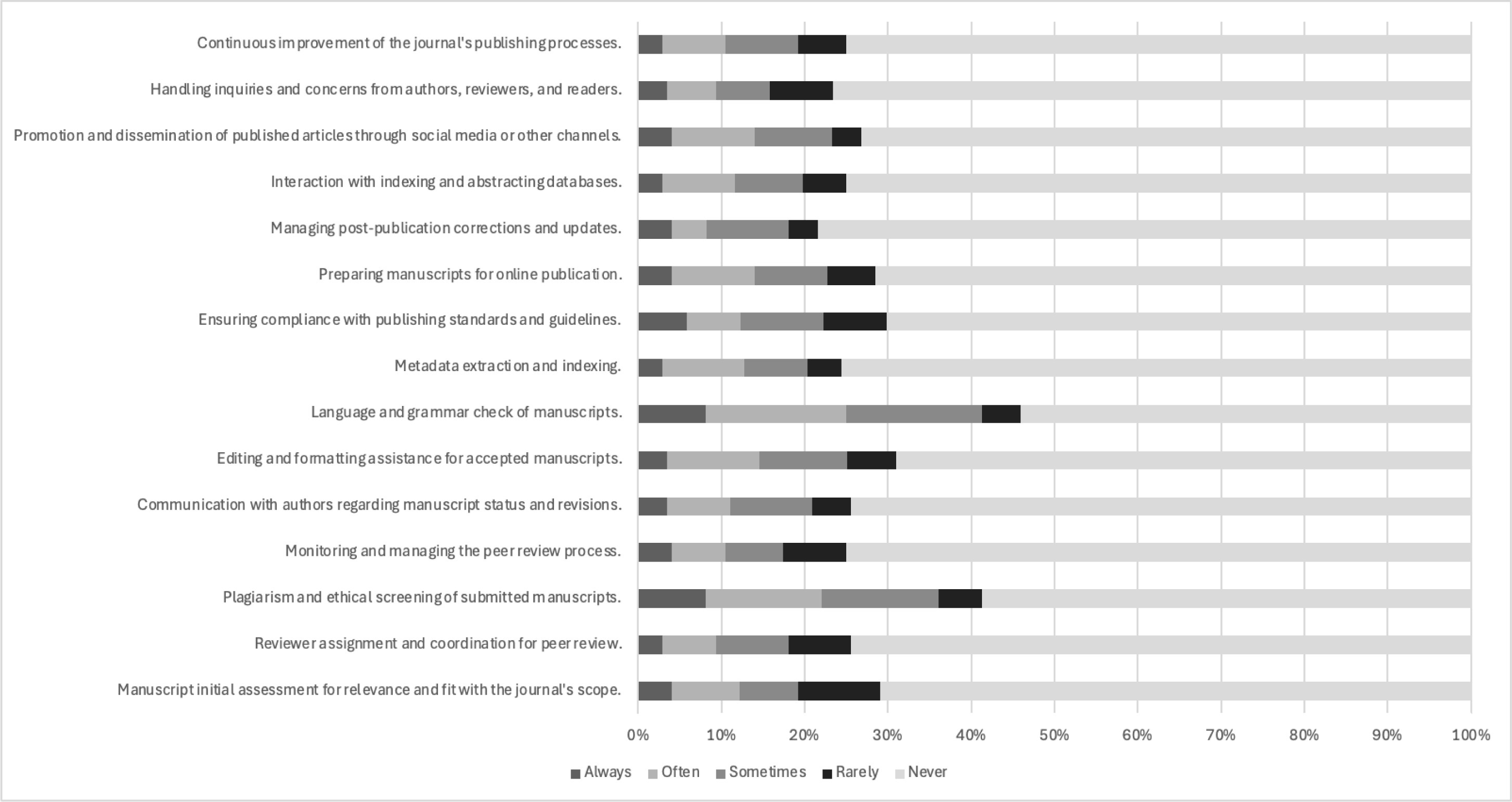
TCIM Editor Frequency of AIC Use in Various Steps of the Scholarly Publishing Process.

Respondents also anticipated a strong role for AICs in the future of scholarly publishing, with 46.0% (n=80/174) rating them as “very important” and 36.8% (n=64/174) as “important” **(Figure 4).** Overall perceptions of AICs’ potential impact were largely positive, with 16.2% (n=28/173) rating the impact as “very positively” and 42.8% (n=74/173) as “positively,” though few also expressed concern, rating the impact as “Negatively” (11.6%) or “very negatively” (8.1%) (**Figure 5**).

**Figure 4.**
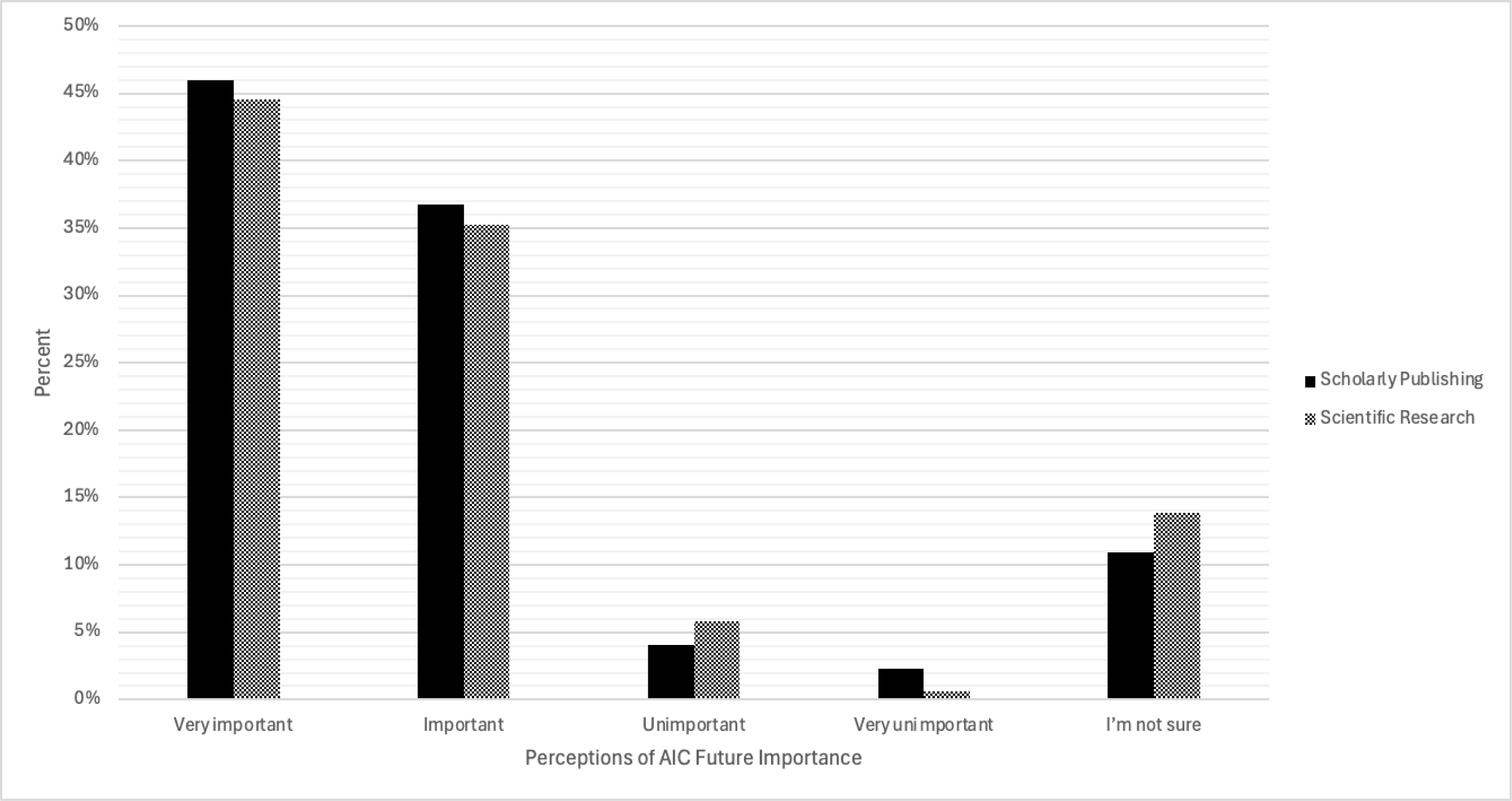
TCIM Editor Perceptions of Future Importance of AICs in Scholarly Publishing and Scientific Research.

**Figure 5.**
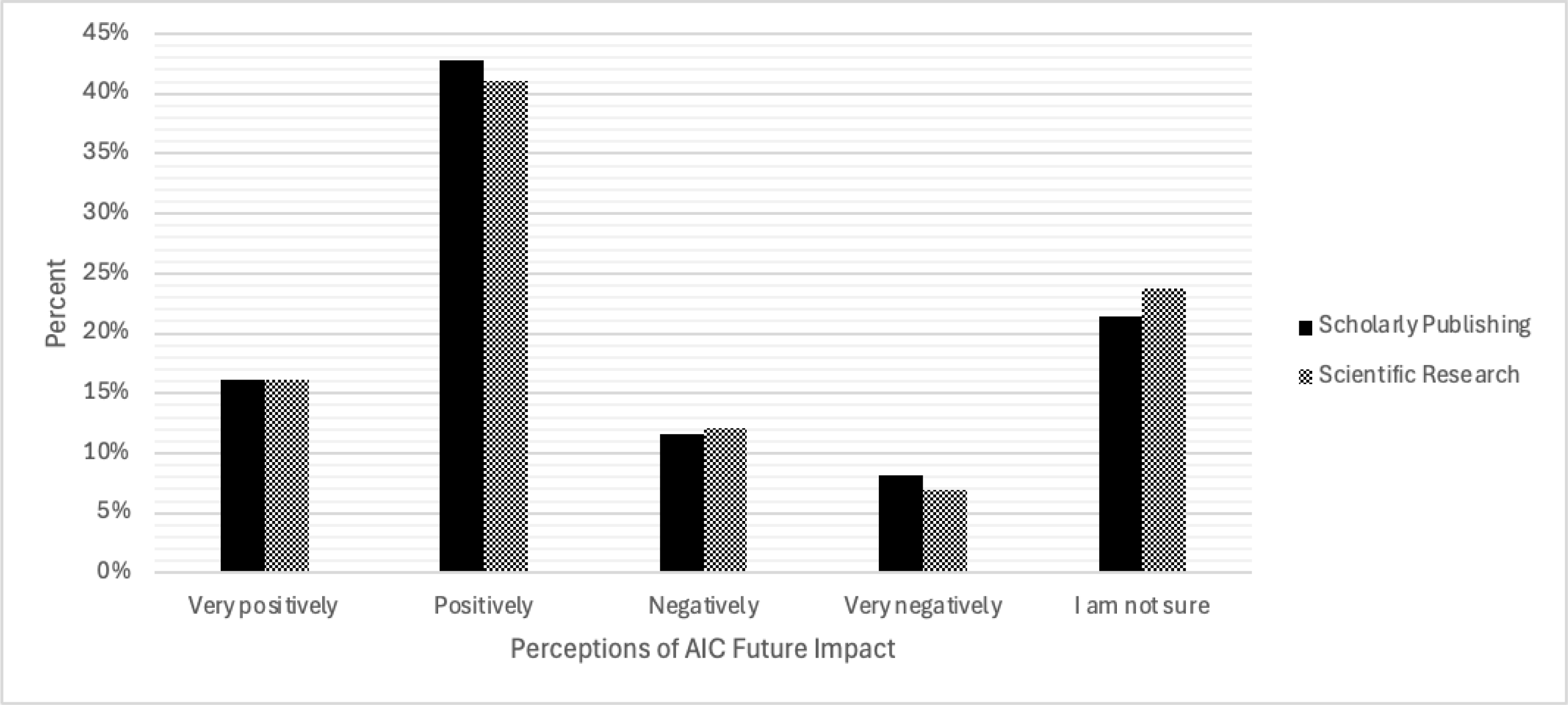
TCIM Editor Perceptions of Potential Future Impact of AICs in Scholarly Publishing and Scientific Research.

### Section 4: Perceived Benefits and Challenges of AI Chatbots in the Scholarly Publishing Process (Q34-36)

Respondents were asked to indicate their level of agreement with a range of proposed benefits associated with using AICs in the editorial and peer review process on a 5-point scale ranging from “strongly disagree” to “strongly agree.” (**Figure 6)**. Overall, agreement levels were high for several core editorial functions. The strongest endorsement (“agree or “strongly agree”) was for reducing the burden on editorial staff by automating repetitive tasks (n=117/167, 70.1%) and offering language and grammar support, assisting authors in improving the quality of their submissions (n=127/167, 76.0%). “Not sure” responses were common across multiple items, particularly for integrating AICs seamlessly with existing editorial tools and systems, enhancing the overall workflow efficiency (n=56/165, 34.0%) and supporting post-publication updates and corrections efficiently, ensuring accuracy of published content (n=52/165, 31.5%).

**Figure 6.**
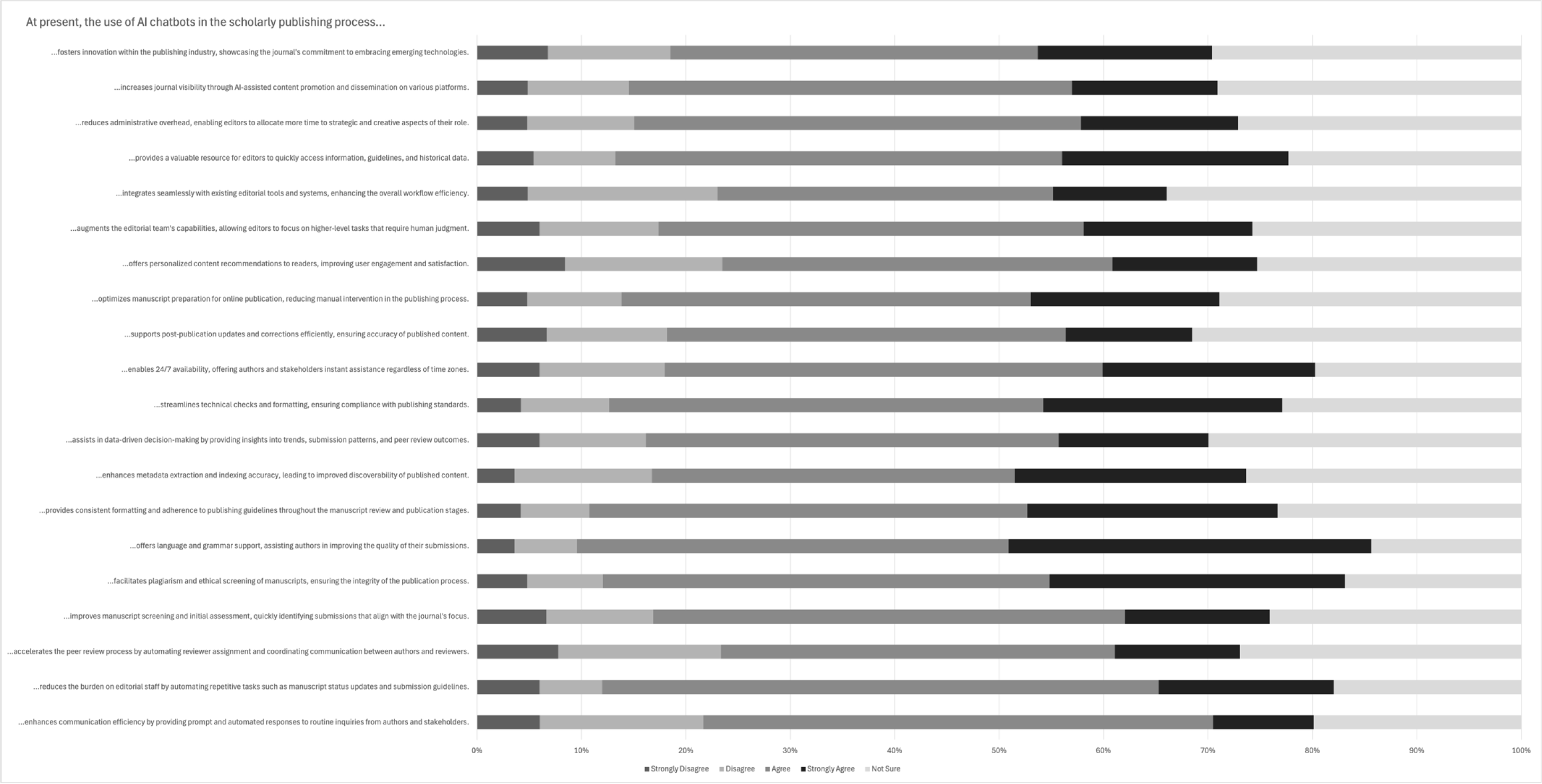
TCIM Editor Perceptions of Proposed Benefits of AICs in the Scholarly Publishing Process.

Respondents were also asked to indicate their level of agreement with various potential challenges to using AICs in the scholarly publishing process (**Figure 7**). There was a particularly strong agreement (“agree” or “strongly agree”) for concerns that AICs use may pose risks in perpetuating biases present in their training data (n = 133/162, 82.1%), lack the human insight necessary to interpret contextual and emotional aspects of interactions with authors and reviewers (n = 130/164, 79.3%), and that AICs present unforeseen ethical quandaries related to accountability, transparency, and responsible AI use (n = 129/163, 79.1%). Respondents recognized practical challenges as well, including the need for additional training and monitoring (n = 128/163, 78.5%), initial investment of time and resources for setup (n = 128/163, 78.5%), and potential difficulties with user acceptance (n = 123/163, 75.5%).

**Figure 7.**
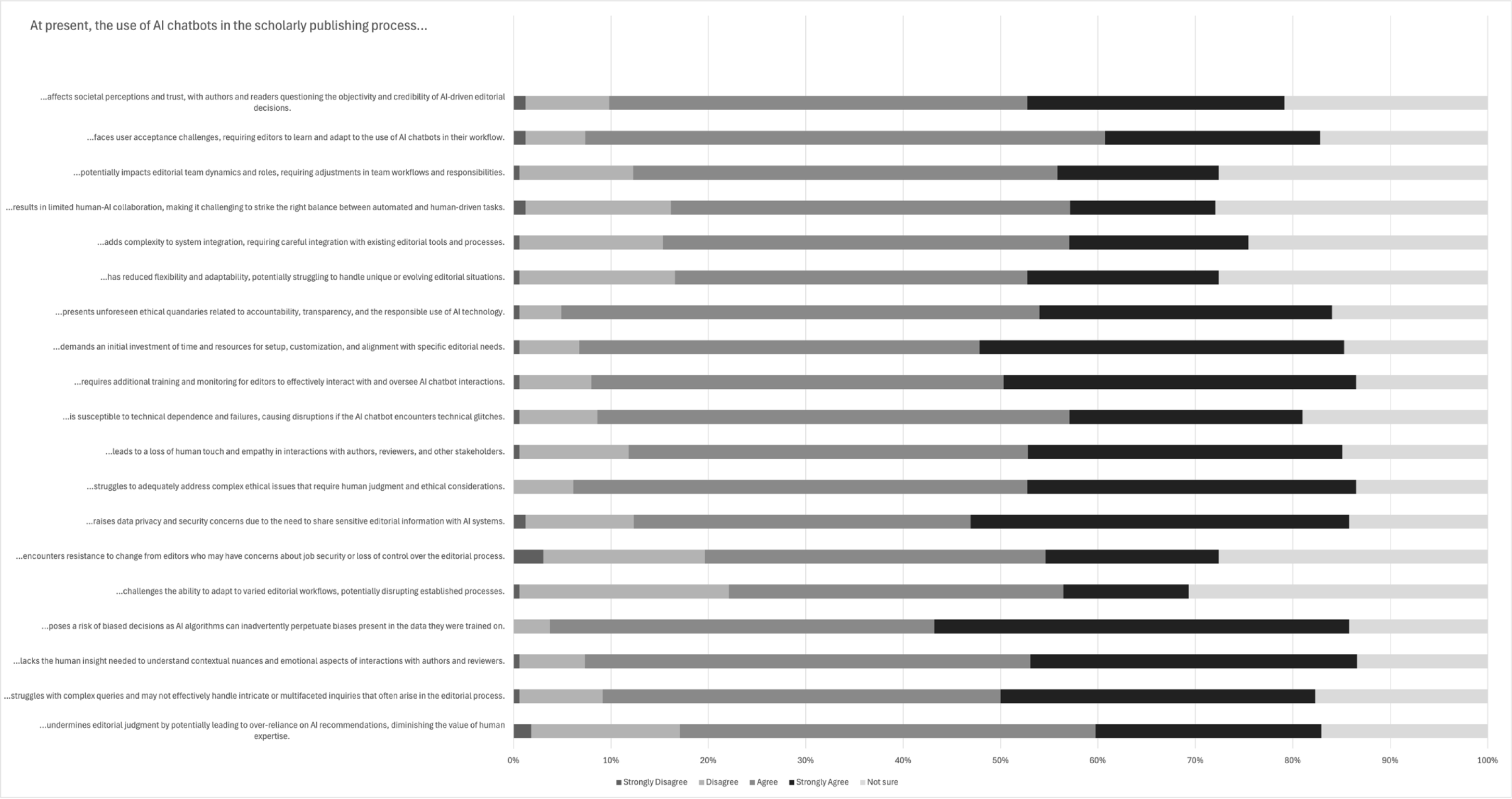
TCIM Editor Perceptions of Proposed Challenges of AICs in the Scholarly Publishing Process.

### Open-Ended Questions

A total of seven open-ended questions offered participants an opportunity to share additional insights and feedback regarding the training and policies made available by affiliated journals or publishers, and the benefits and challenges of AIC use in the editorial and peer review process. The identified themes, subthemes, individual codes, and their frequencies are available in **Appendix 3**, on OSF at https://osf.io/uvfsd. The main themes identified through the thematic analysis include “no AI in authorship or peer review”, “assist with peer review,” “AIC privacy, reliability, and ethical concerns.” Many editors highlighted the value of AICs for discrete, low-risk tasks, particularly language editing, translation, and administrative support, and several reported already using these tools to streamline manuscript review or enhance clarity in peer review reports. Editors also identified potential opportunities for AICs to assist with reference verification, methodological appraisal, and generating ideas for emerging research areas. However, these perceived benefits were consistently tempered by concerns about accuracy, confidentiality, and the risk of over automation eroding the human judgment central to scholarly publishing. Respondents emphasized that hallucinations, bias, and lack of accountability represent threats not only to research quality but also to the integrity of peer review. Policies described by editors reflected this tension: while some journals permit limited use of AICs for grammar and language refinement, many prohibit their use in authorship or peer review and stress the importance of protecting confidential manuscript content. Editors repeatedly underscored the need for clearer guidelines, greater training on responsible and ethical AIC use, and safeguards to preserve the human decision-making essential to rigorous scholarship. Overall, responses reveal a field navigating the balance between leveraging AICs’ practical advantages and maintaining high standards of research quality, transparency, and editorial integrity. Additional details on thematic analysis are available in **Table 4**.

**Table 4:** Thematic Analysis of Open-Ended Survey Responses.

| Thematic Analysis of Medical Journal Editors' Training Modalities for AIC Use Made Available by Journals or Publishers |  |
| --- | --- |
| Themes | Definition and Example |
| Facilitated training | This theme refers to editors' participation in facilitated training opportunities to build skills in using AICs. These include attending online courses and webinars.<br>"Online short course" |
| Responsible Use of AI in Research | This theme refers to editors' training which comprises of ethical and practical considerations when using AICs in research and publishing. These include addressing ethical considerations, navigating biases, and applying AI responsibly in research contexts.<br>"**Familiarity with the system** and how to enhance use of AI to **complement research.**" |
| Thematic Analysis of Medical Journal Editors Regarding Chief Journal/Publisher Policy on AIC use |  |
| No AI in Authorship or Peer Review | This theme refers to policies prohibiting the use of AI in authorship or peer review.<br>"**Do not use AI** for editorial purposes." |
| Permitted For Language/Grammar Use | This theme refers to policies allowing AI use for improving grammar or language clarity.<br>"May be used for **grammar and language improvement** in manuscript preparation only. All uses must be declared." |
| Confidentiality Concerns with AI in Peer Review | This theme refers to policies restricting the use of AI during peer review to protect confidentiality.<br>"“Most of them mention that the content of manuscript **should not be shared** in an AI platform. I only use AI to check the grammar of some sentences.”" |
| Other Permissions/Restrictions on AI use | This theme refers to policies restricting the use of AI during peer review to protect confidentiality.<br>"AI query for background and factual status is **encouraged with caution** as to information provided". |

| Thematic Analysis of Medical Journal Editors' Perceptions of Steps in the TCIM Publishing Process wherein AIC Use Could be Helpful |  |
| --- | --- |
| Manuscript Review | This term refers to the Medical Editors' perspectives that AIC could be helpful with reviewing the language, content, methodologies, data collection, and references of manuscripts.<br>**"Reference screening** (make sure the references are real and that they match up appropriately to the facts being presented in the manuscript)" |
| Assist with Peer Review | This term refers to the Medical Editors' perceptions that AIC could be helpful with the various peer review steps of the TCIM publishing process, including the scoring of reviewer comments in aggregate.<br>**"Just organizing and managing the editorial board"** |
| Research Innovation | This term refers to the Medical Editors' perceptions that AICs could be helpful with identifying and learning about novel topics in emerging fields of TCIM.<br>**"Brain storming** on special issues topics" |
| Publisher Tools | This term refers to the Medical Editors' perceptions that AIC could be helpful with other purposes that help facilitate connections between peer reviewers and manuscript others, such as translation and clarifying comments made by the reviewer.<br>**"FAQ questions regarding how to respond to reviewers recommendations."** |
| Thematic Analysis of Steps in the Scholarly Publishing Process Wherein Medical Journal Editors Have Used AICs |  |
| Manuscript Review | This term refers to the Medical Editors' perspectives that AIC could be helpful with reviewing the language, content, methodologies, data collection, and references of manuscripts.<br>**"Check of correctness and plausibility** of text revised by the authors and assistance in recommending better wording, textflow or identifying missing aspects, in which I do not have enough expertise." |
| Assist with Peer Review | This term refers to the Medical Editors' perceptions that AIC could be helpful with the various peer review steps of the TCIM publishing process, including enhancing reviewer reports. |
|  | “**Translating comments** in English, checking unfamiliar interventions or outcome assessment” |
| **Thematic Analysis of Medical Journal Editors’ Perceptions of Benefits Regarding the Use of AIC in the TCIM Editorial and Peer Review Processes** |  |
| **Supporting the Peer Review Process** | This term refers to the Medical Editors’ perspectives of the most prominent benefit with AIC use being supportive purposes, such as assisting in editing, improving the speed and efficiency of tasks, particularly administrative.<br>“Assist with editorial process as a **supplemental tool** that neither directs nor selects editorial judgements.” |
| **Language Translation and Writing Support** | This term refers to the Medical Editors’ perceptions of other benefits including enhancing the quality of peer review reports and aiding in translation for those editorial board members using English as a second language.<br>“Language conversion” |
| **Thematic Analysis of Medical Journal Editors’ Perceptions of Challenges Regarding the Use of AIC in the TCIM Editorial and Peer Review Processes** |  |
| **Over Automation of Peer Review Process** | This term refers to the Medical Editors’ perceptions of AIC use acting as a potential detriment to the human aspect of publishing due to dependence and overreliance, leading to diminished novel and meaningful research.<br>“The possibility that larger publications will **rely heavily on AI**, removing the Peer Review aspect - suggesting a paper left something out or drew the **wrong conclusions** from the results data set.” |
| **Lack of Accountability/Accuracy with AI Generated Output** | This term refers to the Medical Editors’ perceptions of inaccurate and unverified AIC outputs that may pose a threat of misinformation (i.e., hallucinations) and plagiarism in published content.<br>“ai regularly gives me inaccurate and actually dangerous misinterpretations of the clinical literature.” |
| **Compromised Research Quality Assurance** | In combination with the other two themes, this term refers to the Journal Editors’ perceptions that AIC use may lead to an overall negative impact on the manuscript quality and the general productivity of research in TCIM.<br>“Risk of an increasing number of publications which can be quickly produced but are of LITTLE or NO RELEVANCE. E.g. systematic reviews with searches, |
|  | data extraction and synthesis and even discussion done by AI which **do not address relevant health research questions** or combine data in a meaningless way.” |
| **Thematic Analysis of Medical Journal Editors’ Overall Perceptions of AIC Use in the Scholarly Publishing Process** |  |
| **AIC Privacy, Reliability, and Ethical Concerns** | This term refers to the Medical Editors’ perceptions of Integration, ethical and confidentiality implications with the use of AICs in the scholarly publishing process.<br>“If we **input unpublished papers** into AI, isn't that problematic because then AI can use the content in AI searches?” |
| **Potential Benefits of AICs** | This term refers to the Medical Editors’ perceptions of potential benefits with the use of AIC in the scholarly publishing process.<br>“...I mostly see it at present as being used by authors (**particularly non-native English speakers**) to make their work read more clearly prior to submission.” |
| **Further Training and Guidelines Required** | This term refers to the Medical Editors’ perceptions of the need for editor AIC training with the use of AIC in the scholarly publishing process.<br>“AI should be used very responsibly. It may be useful in certain processes, but overdependence on AI bots may not be advisable. Editors need **a lot of training** for the ethical use of AI.” |
| **Survey Design Feedback** | This term refers to the Medical Editors’ praise and criticism for this study/survey design.<br>**“The survey questions are comprehensive.”** |
| **Implications on Scholarly Process and Human Aspect** | This term refers to the Medical Editors’ perceptions of the implication on scholarly process and human aspect with the use of AIC in the scholarly publishing process.<br>“The **human intelligence** needed in editorial decision making is more of lateral thinking and **beyond algorithmic thinking** of AI tools. Hence AI use should best stay limited to other tasks like suggesting peer reviewers or reminders etc.” |
| **Acceptance with Caution** | This term refers to the Medical Editors’ perceptions of limitations with the use of AIC that need to be addressed before progressing to careful incorporation of these tools in the scholarly publishing process.<br>“AI is **here to stay**, we cannot reverse the history. Journals should instead make sure it is **used** |
|  | **appropriately, and safely.** There always need to be a check of decisions made by AIs.” |
| **General Perspectives on AIC Integration** | This term refers to the Medical Editors’ overall diverse perspectives on AIC integration in the scholarly publishing process.<br>“AI in the world of Editors is a **necessary evil**. We can’t wish it away, given its abilities like time-saving and overcoming language barriers, especially in countries where English is not the first language.” |

## Discussion

The primary objective of this study was to assess the perceptions of editors of TCIM journals regarding the use of AICs in the editorial and peer review processes. In this survey, although most TCIM journal editors were familiar with AICs and many had personal experience using them, over 70% reported that they had never used an AIC for tasks related to their editorial role. Respondents expressed mixed views on their potential application, with strongest support for discrete, text-focused functions, such as language and grammar checks and plagiarism/ethical screening. However, many respondents were less inclined to view AICs as useful for complex, interpretive, or interpersonal editorial tasks, such as managing author communications or handling inquiries. These findings may be explained by several interrelated factors. The integration of AI into scholarly publishing, particularly within TCIM, is newly emerging, with limited field-specific tools tailored to the unique methodological diversity and cultural nuances of TCIM research [21,22]. Editors in TCIM also often navigate heterogeneous evidence bases and culturally embedded knowledge systems, making the automation of complex judgment calls inherently challenging [1,22].

Respondents acknowledged the multiple potential benefits of AICs, particularly in reducing repetitive administrative work, supporting manuscript clarity, and assisting with technical checks. Nonetheless, strong agreement also emerged around key challenges, including the inability of AICs to interpret contextual or culturally embedded nuances, risks of bias in decision-making, and unresolved ethical concerns such as accountability and transparency of prompt outputs. Practical barriers such as the lack of formal training, minimal institutional policies, and uncertainty around integration into existing workflows were also prominent.

Despite limited current use, most respondents anticipated that AICs would be important to the future of scholarly publishing. Nearly all respondents (87.7%) believed that at least some training would be required for effective adoption, yet most reported that their journal or publisher currently did not offer any available AI policies or training opportunities for guiding responsible use. Bridging this gap will require proactive institutional strategies to formulate consistent training initiatives, clear publisher AI policies, and transparency in how AI is evaluated and integrated within the scholarly publishing process [2,3,15].

### Comparative Literature

The integration of AICs into scholarly publishing has garnered attention in recent years for their ability to perform repetitive, time-consuming tasks that place a burden on editorial teams. Kousha & Thelwall, 2023 conducted a review of AI-assisted publishing and peer-review tools, outlining their applications across manuscript screening, plagiarism detection, and reference verification, which collectively streamline early-stage editorial processes [23]. Our findings echo these observations, as TCIM editors were most receptive to AIC use for well-defined, non-critical activities such as grammar editing and ethical compliance screening. This preference suggests that, across publishing contexts, editors tend to support AI applications that produce consistent, verifiable outcomes with minimal interpretive judgment.

Nevertheless, the potential of AICs to contribute meaningfully to more complex editorial activities, such as providing an overall assessment of a manuscript, remains unclear. Existing literature has questioned the reliability of these tools when assessing research quality or detecting fabricated content [24,25]. Dathathri et al., 2024 conducted an experimental study testing watermarking methods across multiple large language models to track AI-generated outputs, demonstrating both the technical feasibility and limitations of current detection systems in maintaining research integrity [24].

A cross-sectional survey of biomedical EiCs conducted by Ng et al., 2025 has comparable findings. Sampling EiCs across five major publishers, most respondents were familiar with AICs (66.7%) but had not used them in their editorial role (83.7%), and many expressed interest in further training (64.4%) [15]. Perceived benefits of AICs clustered around low-risk tasks, language/grammar checks (70.8%) and plagiarism/ethics screening (67.3%), whereas views on accelerating peer review were mixed. Key barriers included setup time/resources (83.7%), training needs (83.9%), and ethical considerations (80.6%). The alignment with our findings in this study suggests that responsible adoption will hinge on scoped use cases, targeted editor training, and clear governance rather than broad automation.

Ng et al., 2025 also conducted two other large-scale international cross-sectional surveys [26, 27]. The first study surveyed 1,209 university students and postdoctoral fellows from over 40 countries across the fields of medicine and life sciences. While most participants rated AICs as ‘important’ or ‘very important’ for future education (87.5%), only 41% believed that the tools provided consistent and reliable information with over 80% expressing concern about misinformation, overreliance, and threats to academic integrity [26]. Complementing this, a second international survey of 2,165 medical and life science researchers found that although 60.5% were familiar with AICs and nearly 45% had used them for purposes relating to the scientific process, only 11% reported institutional training and 10% reported existing policies [27]. Together, these studies illustrate that although academic stakeholders perceive AICs as powerful tools for learning and productivity, persistent concerns remain about transparency, ethical accountability, and overreliance.

Ethical frameworks from organizations such the European Commission underscore the need for transparency, accountability, and disclosure when AI is used in editorial workflows [28]. In particular, the European Commission advises against the substantial use of generative AI in sensitive editorial activities that could influence other researchers or institutions, such as peer review and the evaluation of research proposals [28]. This caution reflects concerns that reliance on such tools in decision-making roles may introduce risks of biased or inaccurate assessments due to known limitations, including hallucinations and algorithmic bias. Additionally, limiting the use of generative AI in these contexts helps protect the confidentiality and integrity of unpublished scholarly work, reducing the likelihood of unintended exposure or incorporation into AI training models. However, the absence of formalized and consistent AI-use policies among TCIM journals and publishers observed in our study suggests that TCIM outlets may fall behind larger or better-resourced publishers in operationalizing these ethical standards.

### Strengths and Limitations

The methodological strengths of this study include a comprehensive approach to data collection and the targeted sampling method. By using a list of unique TCIM journals and manually collecting the names and email addresses of editors, the study ensured a robust and representative sample of the target population. The structured questionnaire, validated through pilot testing, facilitated the collection of detailed and relevant data on editors’ perceptions. Maintaining anonymity by not collecting personal identifiers with the survey may also have encouraged our participants to share their perceptions and declarations (e.g., past use of AICs) with candidacy and openness.

However, the study also has certain limitations. The reliance on self-reported data may introduce response and recall bias, as editors with either a more favourable or more critical viewpoint towards AICs may have been more likely to participate [29]. Additionally, reported beliefs about AI may not fully align with actual patterns of use, as respondents may underreport or overreport engagement with AICs due to social desirability or uncertainty regarding evolving norms around AI adoption. Another limitation is that researchers who do not speak English were excluded because of the language requirements, which may reduce the relevance and transferability of the findings to individuals who predominantly publish in other languages. Similarly, those with limited English proficiency may also face barriers to participating in the survey, further restricting the diversity of perspectives captured. In addition, the response rate may be inaccurately low due to factors such as inactive email accounts. The cross-sectional survey study design is susceptible to recall bias and non-response bias, both of which may influence the accuracy and reliability of the results. Furthermore, the rapid pace of AI tool development represents an additional limitation, as respondents’ perceptions and practices may change quickly over time. Despite these limitations, the study has provided valuable insights into the current state of AI integration in the editorial and peer-review processes of TCIM journal publishing and provides the basis for future research and technological advancements in this field.

## Conclusion

TCIM journal editors recognize the potential of AI chatbots to enhance efficiency of editorial tasks, improve manuscript quality, and support routine editorial functions. However, the actual use of AICs remains limited due to practical barriers, ethical concerns, and a lack of training and policy guidance. Most editors anticipate that AICs will play an important role in the development of the field of TCIM scholarly publishing, underscoring the need for tailored training programs, transparent governance, and culturally sensitive AI tools. The findings of this survey hence may have important implications for the future integration of AICs in TCIM publishing workflows. Understanding editors’ perceptions provides valuable insights into both the acceptance of, and barriers, to AI adoption, informing the development of tools and guidelines that align with the specific needs and values of the TCIM research community. Furthermore, this work contributes to the broader discourse on the ethical use of AI, emphasizing the importance of reinforcing transparency and accountability to promote responsible, effective implementation while maintaining the integrity and rigor of scholarly publishing.

## Data Availability

All relevant materials and data are included in this manuscript or posted on the Open Science Framework.

https://doi.org/10.17605/OSF.IO/PTFXS

## List of Abbreviations

AI: artificial intelligence
CHERRIES: Checklist for Reporting Results of Internet E-Surveys
OSF: Open Science Framework
STROBE: STrengthening the Reporting of OBservational studies in Epidemiology
TCIM: traditional, complementary, and integrative medicine

## Declarations

### Ethics Approval and Consent to Participate

We sought and were granted ethics approval by the University Hospital Tübingen Research Ethics Board prior to beginning this project (REB Number: 081/2025BO2).

### Consent for Publication

All authors consent to this manuscript’s publication.

### Availability of Data and Materials

All relevant materials and data are included in this manuscript or posted on the Open Science Framework: https://doi.org/10.17605/OSF.IO/PTFXS

### Competing Interests

The authors declare that they have no competing interests.

### Funding

This study was unfunded.

## Acknowledgements

We would gratefully acknowledge the survey pilot testers for their time and feedback.

## Authors’ Contributions

JYN: designed and conceptualized the study, collected and analysed data, co-drafted the manuscript, and gave final approval of the version to be published.

DB: collected and analysed data, co-drafted the manuscript, and gave final approval of the version to be published.

MK: collected and analysed data, co-drafted the manuscript, and gave final approval of the version to be published.

ND: collected and analysed data, co-drafted the manuscript, and gave final approval of the version to be published.

DF: collected and analysed data, made critical revisions to the manuscript, and gave final approval of the version to be published.

JWK: collected and analysed data, made critical revisions to the manuscript, and gave final approval of the version to be published.

AK: collected and analysed data, made critical revisions to the manuscript, and gave final approval of the version to be published.

JL: collected and analysed data, made critical revisions to the manuscript, and gave final approval of the version to be published.

AM: collected and analysed data, made critical revisions to the manuscript, and gave final approval of the version to be published.

PO: collected and analysed data, made critical revisions to the manuscript, and gave final approval of the version to be published.

JP: collected and analysed data, made critical revisions to the manuscript, and gave final approval of the version to be published.

IV: collected and analysed data, made critical revisions to the manuscript, and gave final approval of the version to be published.

EY: collected and analysed data, made critical revisions to the manuscript, and gave final approval of the version to be published.

KY: collected and analysed data, made critical revisions to the manuscript, and gave final approval of the version to be published.

AZ: collected and analysed data, made critical revisions to the manuscript, and gave final approval of the version to be published.

JZ: collected and analysed data, made critical revisions to the manuscript, and gave final approval of the version to be published.

MSL: provided methodological guidance, made critical revisions to the manuscript, and gave final approval of the version to be published.

YSL: provided methodological guidance, made critical revisions to the manuscript, and gave final approval of the version to be published.

TMN: provided methodological guidance, made critical revisions to the manuscript, and gave final approval of the version to be published.

TO: provided methodological guidance, made critical revisions to the manuscript, and gave final approval of the version to be published.

CMW: provided methodological guidance, made critical revisions to the manuscript, and gave final approval of the version to be published.

LZ: provided methodological guidance, made critical revisions to the manuscript, and gave final approval of the version to be published.

HC: provided methodological guidance, made critical revisions to the manuscript, and gave final approval of the version to be published.

## Notes

### Competing Interest Statement

The authors have declared no competing interest.

### Clinical Protocols

https://doi.org/10.56986/pim.2025.06.008

### Funding Statement

This study did not receive any funding.

### Author Declarations

We sought and were granted ethics approval by the University Hospital Tubingen Research Ethics Board prior to beginning this project (REB Number: 081/2025BO2).

## References

1. Ng JY, Cramer H, Lee MS, Moher D. Traditional, complementary, and integrative medicine and artificial intelligence: Novel opportunities in healthcare. Integrative Medicine Research. 2024 Mar;13(1):101024. 10.1016/j.imr.2024.101024

2. Roblyer D. AI’s Role in Peer Review in Scholarly Publishing [Internet]. 2024 [cited 2024 Aug 16]. Available from: https://www.highwirepress.com/blog/stm-us-annual-conference-notes-part-3-transforming-the-editorial-and-peer-review-process, https://www.highwirepress.com/blog/stm-us-annual-conference-notes-part-3-transforming-the-editorial-and-peer-review-process/

3. Bauchner H, Rivara FP. Use of artificial intelligence and the future of peer review. Health Aff Sch. 2024 May 3;2(5):qxae058. 10.1093/haschl/qxae058

4. World Health Organization (WHO). [Internet]. Traditional, complementary and integrative medicine; 2023. Available from: https://www.who.int/health-topics/traditional-complementary-and-integrative-medicine

5. Veziari Y, Leach MJ, Kumar S. Barriers to the conduct and application of research in complementary and alternative medicine: A systematic review. BMC Complement Altern Med. 2017 Dec;17(1):166. 10.1186/s12906-017-1660-0

6. Veziari Y, Kumar S, Leach M. Addressing barriers to the conduct and application of research in complementary and alternative medicine: A scoping review. BMC Complement Med Ther. 2021 Dec;21(1):201. 10.1186/s12906-021-03371-6

7. Liu X, Gong T. Artificial intelligence and evidence-based research will promote the development of traditional medicine. Acupuncture and Herbal Medicine. 2024 Mar;4(1):134–5. 10.1097/hm9.0000000000000100

8. Cramer H. Artificial Intelligence, Complementary and integrative medicine: A paradigm shift in health care delivery and research? Journal of Integrative and Complementary Medicine. 2023 Mar 1;29(3):131–3. 10.1089/jicm.2023.0040

9. Chubb J, Cowling P, Reed D. Speeding up to keep up: Exploring the use of AI in the research process. AI Soc. 2022;37(4):1439–57. 10.1007/s00146-021-01259-0

10. Thompson AD, Schmidt-Crawford DA, Lindstrom DL. Challenges for journal editors: An assist from AI? Journal of Digital Learning in Teacher Education. 2024 Apr 2;40(2):74–5. 10.1080/21532974.2024.2348923

11. Fabiano N, Gupta A, Bhambra N, Luu B, Wong S, Maaz M, et al. How to optimize the systematic review process using AI tools. JCPP Adv. 2024 Apr 23;4(2):e12234. 10.1002/jcv2.12234

12. Checco A, Bracciale L, Loreti P, Pinfield S, Bianchi G. AI-assisted peer review. Humanit Soc Sci Commun. 2021 Jan 25;8(1):1–11. 10.1057/s41599-020-00703-8

13. Brown C, Nazeer R, Gibbs A, Le Page P, Mitchell AR. Breaking bias: The role of artificial intelligence in improving clinical decision-making. Cureus. 15(3):e36415. 10.7759/cureus.36415

14. Ng JY, Bhavsar D, Dhanvanthry N, Lee MS, Lee Y-S, Nesari TM, et al. Artificial Intelligence in the editorial and peer review process: A protocol for a cross-sectional survey of traditional, complementary, and integrative medicine journal editors’ perceptions. Perspectives on Integrative Medicine 2025;4:121–4. 10.56986/pim.2025.06.008

15. Ng JY, Krishnamurthy M, Deol G, Al-Khafaji WA-Z, Balaji V, Abebe M, et al. Attitudes and perceptions of biomedical journal editors in chief towards the use of artificial intelligence chatbots in the Scholarly Publishing Process: A cross-sectional survey. Research Integrity and Peer Review 2025;10. 10.1186/s41073-025-00178-8

16. SurveyMonkey [Internet]. [cited 2025 Aug 1]. Available from: https://www.surveymonkey.com/

17. Thomas DR. A general inductive approach for analyzing qualitative evaluation Data. American Journal of Evaluation. 2006 Jun;27(2):237–46. 10.1177/1098214005283748

18. Joffe H, Yardley L. Content and thematic analysis. In: Marks DF, Yardley L, editors. Research Methods for Clinical and Health Psychology. London: SAGE Publications; 2003. p. 56–68. 10.4135/9781849209793.n4

19. Eysenbach G. Improving the Quality of web surveys: The checklist for reporting results of internet e-surveys (CHERRIES). Journal of Medical Internet Research. 2004 Sep 29;6(3):e34. 10.2196/jmir.6.3.e34

20. Von Elm E, Altman DG, Egger M, Pocock SJ, Gøtzsche PC, Vandenbroucke JP. The Strengthening the Reporting of Observational Studies in Epidemiology (STROBE) statement: Guidelines for reporting observational studies. The Lancet. 2007 Oct 20;370(9596):1453–7. 10.1016/S0140-6736(07)61602-X

21. Barić H, Škorić L, Kalanj Bognar S. Editors’ role in shaping the publishing environment and guiding authors in the era of artificial intelligence. Croatian Medical Journal. 2024 Dec;65(6):471–2. 10.3325/cmj.2024.65.471

22. Raja M, Cramer H, Lee MS, Wieland LS, Ng JY. Addressing the challenges of traditional, complementary, and integrative medicine research: An international perspective and proposed strategies moving forward. Perspectives on Integrative Medicine. 2024 Jun 30;3(2):86–97. 10.56986/pim.2024.06.004

23. Kousha K, Thelwall M. Artificial Intelligence to support publishing and peer review: A summary and review. Learned Publishing. 2023 Aug 8;37(1):4–12. 10.1002/leap.1570

24. Dathathri S, See A, Ghaisas S, Huang P-S, McAdam R, Welbl J, et al. Scalable watermarking for identifying large language model outputs. Nature. 2024 Oct 23;634(8035):818–23. 10.1038/s41586-024-08025-4

25. Chauhan C, Currie G. The impact of generative artificial intelligence on research integrity in scholarly publishing. The American Journal of Pathology. 2024 Dec;194(12):2234–8. 10.1016/j.ajpath.2024.10.001

26. Ng JY, Shah AQ, Roni E, Asna M, Brar J, Kathirkamanathan S, et al. Attitudes of medical and life sciences university students and postdoctoral fellows toward AI chatbots in education: an international cross-sectional survey. Scientific Reports. 2026. 10.1038/s41598-026-42085-y

27. Ng JY, Maduranayagam SG, Suthakar N, Li A, Lokker C, Iorio A, et al. Attitudes and perceptions of medical researchers towards the use of artificial intelligence chatbots in the scientific process: An international cross-sectional survey. The Lancet Digital Health. 2025 Jan;7(1). 10.1016/s2589-7500(24)00202-4

28. Guidelines on the responsible use of generative AI in research developed by the European Research Area Forum - European Commission [Internet]. 2024 [cited 2025 Aug 1]. Available from: https://research-and-innovation.ec.europa.eu/news/all-research-and-innovation-news/guidelines-responsible-use-generative-ai-research-developed-european-research-area-forum-2024-03-20_en

29. Wetzel E, Böhnke JR, Brown A. Response biases. The ITC International Handbook of Testing and Assessment. 2016 Jun;349–63. 10.1093/med:psych/9780199356942.003.0024

